# Narrow Is Slow: CT-Derived Anatomic Isthmus 3 Width and Arrhythmogenic Substrate in Repaired Tetralogy of Fallot

**DOI:** 10.64898/2026.10.16.26365751

**Authors:** Marinos Kosmopoulos, Steven Liskov, Rawan Amir, Ronald D Berger, Ari Cedars, Omar Chehab, Jonathan Chrispin, Stacy Fisher, Stephanie Fuller, Hannah Fraint, Aimee K Armstrong, Ilias Ziakas, Konstantinos N Aronis

**Affiliations:** Section of Cardiac Electrophysiology, Division of Cardiology, Department of Medicine, Johns Hopkins University Hospital, Baltimore, Maryland, USA; Division of Cardiology, Department of Medicine, Johns Hopkins University School of Medicine, Baltimore, Maryland, USA; Division of Cardiology, Main Line Health Lankenau Medical Center, Wynewood, Pennsylvania, USA; Adult Congenital Heart Disease Clinic, Department of Cardiovascular Medicine, Mayo Clinic, Rochester, Minnesota, USA; Division of Pediatric Cardiology, Department of Pediatrics, Johns Hopkins School of Medicine, Baltimore, Maryland, USA; Division of Cardiology, Columbia University Medical Center, New York, New York, USA; Division of Pediatric Cardiac Surgery, Department of Surgery, Johns Hopkins School of Medicine, Baltimore, Maryland, USA

**Keywords:** **Key Words:** Tetralogy of Fallot, Adult Congenital Heart Disease, SCAI 3, Ventricular Tachycardia, Cardiac CT

## Abstract

**Background:** Anatomic isthmus 3 (AI 3) is defined as the myocardium between the pulmonary valve annulus and the ventricular septal defect patch that persists after tetralogy of Fallot (ToF) repair. Ventricular tachycardia (VT) propagated through slow conducting AI 3 (SCAI 3) represents a significant cause of morbidity in ToF. There is an unmet need for non-invasive identification of SCAI 3 in patients with ToF.

**Objectives:** To assess whether the anatomic features of AI 3 as derived from cardiac computed tomography (CT) can predict slow conduction velocity during EP study.

**Methods:** This retrospective single-center study included patients with ToF who underwent an electrophysiologic study and had AI 3 width measured from the pre-procedure cardiac CT. All patients underwent programmed ventricular stimulation, and electroanatomic. Logistic regression was performed to assess the relationship between AI 3 width and the presence of SCAI 3.

**Results:** Of 27 consecutive patients with ToF, twelve patients were found to have SCAI 3, and ten patients were found to have SCAI 3 mediated VT. The AI 3 was significantly narrower in patients with SCAI 3 compared to those with normal AI 3 conduction (18.4 ± 6.6 mm vs. 29.2 ± 9.0 mm; p = 0.001). An AI 3 width above 21.5 mm carried a negative predictive value of 80.0%.

**Conclusion:** In adults with repaired ToF referred for electrophysiology study, a narrower CT-derived AI 3 width was associated with SCAI 3 identification. AI 3 width measured on a CT obtained before PV replacement could help guide invasive risk stratification.

## Introduction

Tetralogy of Fallot (ToF) is the most common cyanotic congenital heart defect. With surgical repair most patients survive into adulthood.^1,2^ This growing population is at significant risk for late ventricular arrhythmia: the 30-year prevalence of sustained ventricular tachycardia (VT) approaches 14%, and sudden cardiac death accounts for up to one-third of late mortality.^1,3–5^. Surgical correction creates zones of inexcitable tissue (scarring or patches) in the right ventricle that form the borders of anatomic isthmuses (AI) composed of excitable viable myocardium.^6^ With chronic right ventricular remodeling from volume overload, these AIs remodel electrophysiologically, with slowing of the conduction velocity, creating Slowly Conducting Anatomical Isthmuses (SCAI). Four AIs have been described in patients with rTOF.^7,8^ AI 3 (the anatomic isthmus between the pulmonary valve annulus and the ventricular septal defect (VSD) patch) is universally present in patients with rTOF. When slow conduction (SCAI 3) develops, it is the dominant substrate for VT in this population.^7^

The clinical relevance of AI 3 is further amplified by its position immediately beneath the native pulmonary valve (PV) annulus, which places the isthmus directly in the path of PV intervention. Pulmonary regurgitation is an expected long-term sequela of ToF repair, with a prevalence ranging from 40% to 85%, and progressive right ventricular dilation resulting from chronic volume overload frequently necessitates PV replacement.^9^ Transcatheter PV replacement either covers or obscures intracardiac access to AI 3, rendering it permanently inaccessible to catheter ablation. Identifying which patients harbor a SCAI 3 and are therefore at risk for sustained SCAI 3-mediated VT before PV replacement is therefore critical. This currently requires an invasive electrophysiology study (EPS) with assessment of VT inducibility.^10^ A non-invasive method to identify at-risk patients before EPS remains an unmet need.

Cardiac computed tomography (CT) identifies the surgical anatomy that bounds the anatomic isthmuses, including the VSD patch, ventriculotomy, and RVOT, which are commonly demarcated by calcification. CT-derived wall thickness at these isthmuses tracks their functional conduction properties, with thinner myocardium corresponding to slower conduction and block^8^. Whether AI 3 width on CT is associated with VT inducibility has not been investigated.

In the present study, we examined whether the transverse width of AI 3, measured on pre-procedural cardiac CT, was associated with the presence of SCAI 3 and the inducibility of SCAI 3-dependent VT during electrophysiology studies in adults with rToF.

## Methods

### Study Design

This was a retrospective, single-center observational study enrolling adult patients (>18 years) referred to the Johns Hopkins Adult Congenital Electrophysiology Program. This study was approved by the Johns Hopkins Medicine Institutional Review Board. Informed consent was waived given the retrospective design of the study. The study followed the Declaration of Helsinki and is reported in accordance with the STROBE guidelines^11^.

### Patient Population

Patients were identified by querying the Adult Congenital Heart Disease Electrophysiology registry. The inclusion criteria were (1) age ≥18 years; (2) rToF; (3) an electrophysiology study (EPS) with right ventricular electroanatomical mapping and (4) a pre-procedural contrast-enhanced cardiac CT available within six months from the EPS. The collected baseline characteristics are reported in the study supplement.

### Pre-procedural CT

Each patient underwent pre-procedural cardiac computed tomography (CT) within six months of the index procedure. Cardiac-gated CT angiography was performed on a 320-slice scanner (Aquilion ONE, Canon Medical Systems) with a gantry rotation time of 275 ms and 0.5-mm slice thickness. Prospective or retrospective ECG gating was used; dynamic ventricular function was not assessed. Images were obtained in the arterial (first-pass) phase following intravenous administration of iodinated contrast, and iterative three-dimensional reconstructions of the cardiac chambers were generated at 75% of the R-R interval. Tube voltage ranged from 80 to 120 kV and tube current from 390 to 680 mA across studies; in a subset, automated tube-current and tube-voltage modulation were applied. The acquisition was optimized for the cardiac chambers and great vessels rather than for the coronary arteries.

### CT Post-Processing and AI 3 Assessment on CT

CTs were processed using ADAS 3D imaging software (version 2; ADAS3D Medical, Barcelona, Spain). The RV borders were identified using standard anatomical boundaries from the 3D image reconstruction, identifying the tricuspid annulus, atrioventricular groove, interventricular septum, and pulmonary artery, as previously described.^12–14^ The software generated automated boundaries of the RV, which could be manually adjusted by the study investigators (MK and SL). A parametric map of the RV wall thickness was created using a semi-automatic method: the operator adjusted the epicardial boundary of the RV wall, and the software provided a quantitative assessment of the wall thickness between the endocardial and epicardial surfaces. To standardize the RVOT segmentation, the RVOT interventricular septum thickness was assigned entirely to the RV. Epicardial fat was excluded from the measurement of the

RV wall thickness. Cardiac CTs were analyzed by two cardiologists with training in cardiac CT (MK and SL), each blinded to the other’s measurements and EPS outcomes. The AI 3 width was defined as the geodesic distance on the 3D model between the part from the pulmonary orifice and the VSD patch that had wall thickness > 1 mm. Tissue with a thickness of 1 mm or less was considered the border of the isthmus, demarcating the VSD patch, pulmonary trunk, and non-excitable scarred right ventricular myocardium. The 1-mm threshold was specified a priori, before any measurement was performed, based on published multidetector CT data in repaired ToF in which anatomic isthmuses with documented conduction block were substantially thicker than 1 mm (median 2.6 mm, first quartile 2.1 mm).^8^. The thickness border does not require visible patch calcification, the usual CT landmark, which was present in only 46 of 61 patients (75%) in that series [8], and therefore applies even when calcification is absent. Third, 1 mm approximates twice the 0.5-mm slice thickness of our acquisitions, near the limit of reliable resolution. A low threshold is also conservative, since retaining marginal tissue widens the measured isthmus and biases the observed association toward the null (Supplemental Figures S2, S3). The threshold was applied unchanged to every measurement; its derivation, coincidence with a native interval boundary of the ADAS 3D thickness map, and the proportion of isthmus surface it excludes are detailed in the Supplemental Methods and Results (**Supplemental Figures S2 and S3**). The inter-reader reproducibility of the AI 3 width measurement was assessed as a subset of the first 10 consecutive patients.

### Electrophysiology Study and 3D Mapping

All procedures were performed under general anesthesia. Programmed ventricular stimulation (PVS) was performed from the right ventricular (RV) apex and the RV outflow tract at three drive cycle lengths (500, 400, and 350 ms); a single extrastimulus was used to determine the ventricular effective refractory period (VERP) for each drive train, and double and triple extrastimuli were then introduced beginning 60 ms above the VERP and were decremented to refractoriness or to a minimum coupling interval of 180 ms. The protocol was performed at baseline and repeated during isoproterenol infusion, titrated to a heart rate above 110 beats/min. Inducible VT was defined as sustained monomorphic VT lasting at least 30 seconds or causing hemodynamic compromise.

Omnipolar voltage and activation maps were acquired in sinus rhythm with the EnSite X mapping system (Abbott) and a multielectrode Advisor HD Grid catheter (Abbott), with low-voltage tissue defined as an omnipolar voltage below 1.0 mV and dense scar below 0.1 mV. When the sinus activation wavefront converged on AI 3, activation mapping of the isthmus was repeated during pacing lateral and/or septal to AI 3 to unmask potentially inapparent conduction slowing. An AI 3 with a conduction velocity below 0.5 m/s was defined as slowly conducting AI 3 (SCAI 3). A VT was classified as SCAI 3-dependent if at least one of the following criteria was met: activation mapping showing diastolic potentials within SCAI 3, entrainment mapping showing an in-circuit response from SCAI 3, or a VT exit site localized to SCAI 3 by pace mapping. In patients who underwent ablation, radiofrequency energy was delivered with a contact-force–sensing TactiFlex catheter (Abbott) at 35 W, with ablation endpoints of VT non-inducibility, bidirectional block across SCAI 3, and loss of capture during pacing at 10 mA and 3 ms, across the ablation line.

### Outcomes

The primary outcome was the presence of SCAI 3 during invasive EP mapping, as defined above. The primary exposure was the AI 3 width, measured using pre-procedural cardiac CT. Secondary outcomes were (1) the association between AI 3 width and inducibility of SCAI 3-mediated VT, (2) inter-reader reproducibility of the CT AI 3 width measurement, and (3) clinical events during follow-up. Clinical events were ascertained from the medical record through the most recent available follow-up (up to one year after the index EPS) and comprised death, sustained VT, appropriate device therapy, and repeat VT ablation. Because of the small number of events and the limited follow-up, clinical events were summarized descriptively and were not used to assess the prognostic value of AI 3 width. Exploratory outcomes included the associations of SCAI 3 width and SCAI 3-dependent VT inducibility with tachycardia cycle length and age at EPS, age at ToF repair, and time since surgical repair.

## Statistical Analysis

Continuous variables were summarized as mean ± SD or median [IQR] according to their distribution, and categorical variables as counts (percentages). The distribution of continuous variables was assessed using the Shapiro–Wilk test. Comparisons between patients with and without SCAI 3-dependent VT were performed using the independent-samples t-test for normally distributed continuous variables, the Wilcoxon rank-sum test for non-normally distributed continuous variables, and the chi-square test or Fisher’s exact test when expected cell counts were fewer than five, for categorical variables.

To evaluate the association between CT-derived AI 3 width and the presence of SCAI 3 or SCAI 3-dependent VT inducibility, we used univariable logistic regression, with results expressed as odds ratios and 95% confidence intervals. The discriminative performance of AI 3 width was assessed using receiver operating characteristic (ROC) analysis; the optimal cut-point was identified using the Youden index. Sensitivity, specificity, and negative and positive predictive values were calculated at this threshold. Internal validation was performed using bootstrap resampling (B = 1,000 resamples) to estimate optimism-corrected model performance, following the approach described by Harrell et al.^15^ In each resample, the model was refit on the bootstrap sample and evaluated both on that sample (training performance) and on the original dataset (test performance). The average difference between training and test performance across resamples (the optimism) was subtracted from the apparent performance of the original model to obtain optimism-corrected estimates of discrimination (C-statistic), calibration (slope and intercept), and overall performance (Nagelkerke R² and Brier score).^16^ Analyses were conducted in R using the validate() function from the rms package.^17^

The inter-reader reproducibility of the AI 3 width measurement was assessed using (a) the intraclass correlation coefficient reported with its 95% confidence interval and (b) Bland–Altman analysis reporting the mean bias and 95% limits of agreement. Clinical follow-up events were summarized descriptively; the small number of events precluded time-to-event modeling.

Exploratory analyses assessed the relationship between SCAI 3 width and tachycardia cycle length and time since surgical repair. Given the small number of observations, associations were first quantified with the Spearman rank correlation coefficient, followed by linear regression to estimate the magnitude of the association, reporting the coefficient of determination (R²) and p-value. Influential observations were identified using Cook’s distance, and a sensitivity analysis was performed excluding observations with a Cook’s distance exceeding 1. The association between age at EPS, age at ToF repair, and time since surgical repair with the presence of SCAI 3 and SCAI 3-dependent VT inducibility was assessed using univariable logistic regression. Given the limited number of outcome events, no multivariable analysis was performed. All tests were two-sided, and p < 0.05 was considered statistically significant. Analyses were performed in R version 4.4.1 (R Core Team, Vienna, Austria).

## Results

### Baseline Characteristics

Between February 2022 and February 2026, 29 adults with repaired ToF were referred for electrophysiology study and possible VT ablation. Two were excluded: one had atrial flutter and did not undergo ventricular mapping, and one had inadequate right ventricular opacification on CT. The analytic cohort therefore comprised 27 patients, each with a measurable AI 3 width on pre-procedural CT and a measured AI 3 conduction velocity (Supplemental Figure S1). SCAI 3 was identified in 12 patients, and 15 had AI 3 without slow conduction; these two groups formed the primary analysis cohort. Among the 12 patients with SCAI 3, SCAI 3-dependent VT was inducible in 10 and not inducible in 2. The secondary analysis therefore compared the 10 patients with inducible SCAI 3-dependent VT against the remaining 17 (15 without SCAI 3 and 2 with SCAI 3 in whom SCAI 3-dependent VT was not inducible).

The baseline characteristics of the primary cohort are summarized in **Table 1**. The median age was 37 years (28.0–50.0) and 40.7% were men. The mean AI 3 width was 24.4 ± 9.6 mm. Twelve patients had inducible VT at the time of EPS. Of these, ten had a VT exit site localized to SCAI 3; one patient had SCAI 2-dependent VT and one SCAI 1-dependent VT. Patients with SCAI 3 were older and more likely to have had a history of VT and a prior implantable cardioverter-defibrillator (ICD) implantation. No significant differences were observed in RV function or baseline ECG parameters between the two groups. Patients with SCAI 3 were more likely to have undergone EP study for VT ablation, while patients with normal AI 3 velocity were more likely to be referred for assessment of palpitations or syncope.

**Table 1:** Baseline characteristics in patients with and without SCAI 3 VT.

| Variable | Study Population (N= 27) | SCAI 3 (N = 12) | Normal AI 3 velocity (N=15) | p-values |
| --- | --- | --- | --- | --- |
| Age at EP study (years) | **37 [28, 50]** | **50.0 [41.8, 57.3]** | **28.0 [25.5, 36.5]** | **0.002** |
| Male Gender (%) | 11(40.7) | 6(50) | 5(33.3) | 0.630 |
| Race (% White) | 18 (66.7) | 8 (66.7) | 10 (66.7) | 1.000 |
| Anatomic Isthmus 3 Width | **24.4 ± 9.6** | **18.4±6.6** | **29.2 + 9.0** | **0.001** |
| Number of thoracotomies | 2[1,2] | 2[2,2.3] | 2[1,2] | 0.107 |
| Age at TOF repair (years) | 2[1,3.5] | 3[1, 4.3] | 2[1,3] | 0.232 |
| BMI (kg/m<sup>2</sup>) | 27.5 [24.3, 32.7] | **31.5 [26.9, 34.0]** | **27.3[21.2, 28.4]** | **0.045** |
| Baseline QRS duration (ms) | 160[146, 176] | 168[146, 188] | 156[146, 172] | 0.196 |
| Baseline severe QRS fragmentation (%) | 18 (66.7) | 8(66.7) | 10(66.7) | 1.000 |
| Baseline RBBB (%) | 21(77.8) | 8(66.7) | 13(86.7) | 0.438 |
| VT history | **7 (25.9)** | **6 (50.0)** | **1 (6.7)** | **0.035** |
| ICD before EP study (%) | **5 (18.5)** | **5 (41.6)** | **0 (0)** | **0.023** |
| AAD before EP study | 12 (44.4) | 7 (58.3) | 5 (33.3) | 0.194 |
| RV dysfunction (%) | 10 (37.0) | 6 (50.0) | 4 (26.7) | 0.212 |
| RV basal diameter (cm) | 5.2 [4.3- 5.9] | 5.3 [4.3- 6.6] | 5.0 [4.6- 5.6] | 0.978 |
| RVSP (mm Hg) | 32 [27.0, 59.0] | 39 [28, 65.3] | 32 [27, 41.5] | 0.617 |
| Pulmonic Valve replacement before study (%) | 9 (33.3) | 6 (50.0) | 3 (20.0) | 0.219 |
| EP study indication: Pre Pulmonic Valve replacement (%) | 18 (66.7) | 7 (58.3) | 11 (73.3) | 0.411 |
| EP study indication: Palpitations/Syncope | 4 (14.8) | 0 (0) | 4 (26.7) | 0.053 |
| EP study indication: Clinical VT | **5 (18.5)** | **5 (41.7)** | **0 (0.0)** | **0.006** |

EP: Electrophysiology, BMI: Body mass index, RBBB: Right Bundle Branch Block, VT: Ventricular Tachycardia, ICD: Intracardiac Defibrillator-Cardioverter, AAD: Antiarrhythmic drug before EP study, RV: Right Ventricle, RVSP: Right Ventricular Systolic Pressure. Normally distributed continuous variables are reported as mean ± standard deviation. Not normally distributed variables are reported as median [25,75% IQR]. Bold font indicates statistical significance.

### Narrower AI 3 by CT is Associated with the presence of SCAI 3

Patients with SCAI 3 had significantly shorter isthmuses than those with normal AI 3 velocities (18.4 ± 6.6 mm vs. 29.2 ± 9.0 mm; p = 0.001; **Figure 1**). Univariable logistic regression demonstrated a significant association between isthmus width and SCAI 3 identification in electroanatomic mapping. Each 5 mm decrease in width was associated with 2.31-fold higher odds for the presence of SCAI 3 (95% CI 1.21–4.40, p = 0.011). Apparent discrimination was good, as demonstrated in the ROC analysis (**Figure 1B**, c = 0.822 (95% CI 0.664–0.980); Somers’ Dxy = 0.644; Nagelkerke R² = 0.420; Brier score 0.171). A threshold of 21.5 mm (predicted probability 0.52), yielded a sensitivity of 75.0% and specificity of 80.0%, with positive and negative predictive values of 75.0% and 80.0% and an overall accuracy of 77.8%. Representative examples are shown in **Figure 2**.

**Figure 1.**
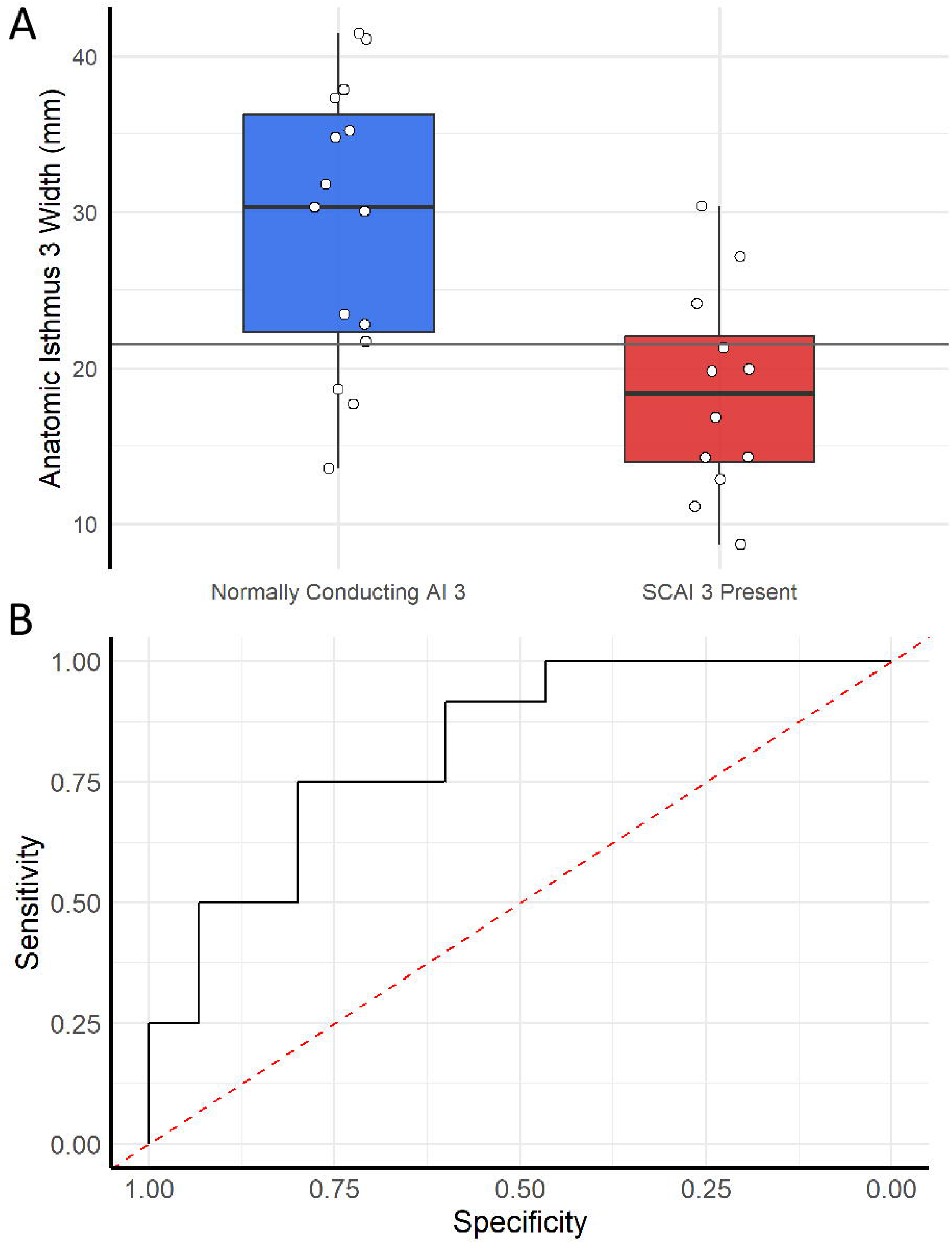
**A.** Box-and-whiskers plot demonstrating the Isthmus 3 width in patients with (red bar) and without (blue) slow conducting anatomic isthmus 3. The grey solid line corresponds to the derived threshold of 21.5 mm as the cutoff between patients who have SCA3 and those who have AI 3 without slow conduction. **B.** Receiver operating characteristic (ROC) curve for AI 3 isthmus width and identification of SCAI 3 during EPS. The area under the curve was 0.822.

**Figure 2.**
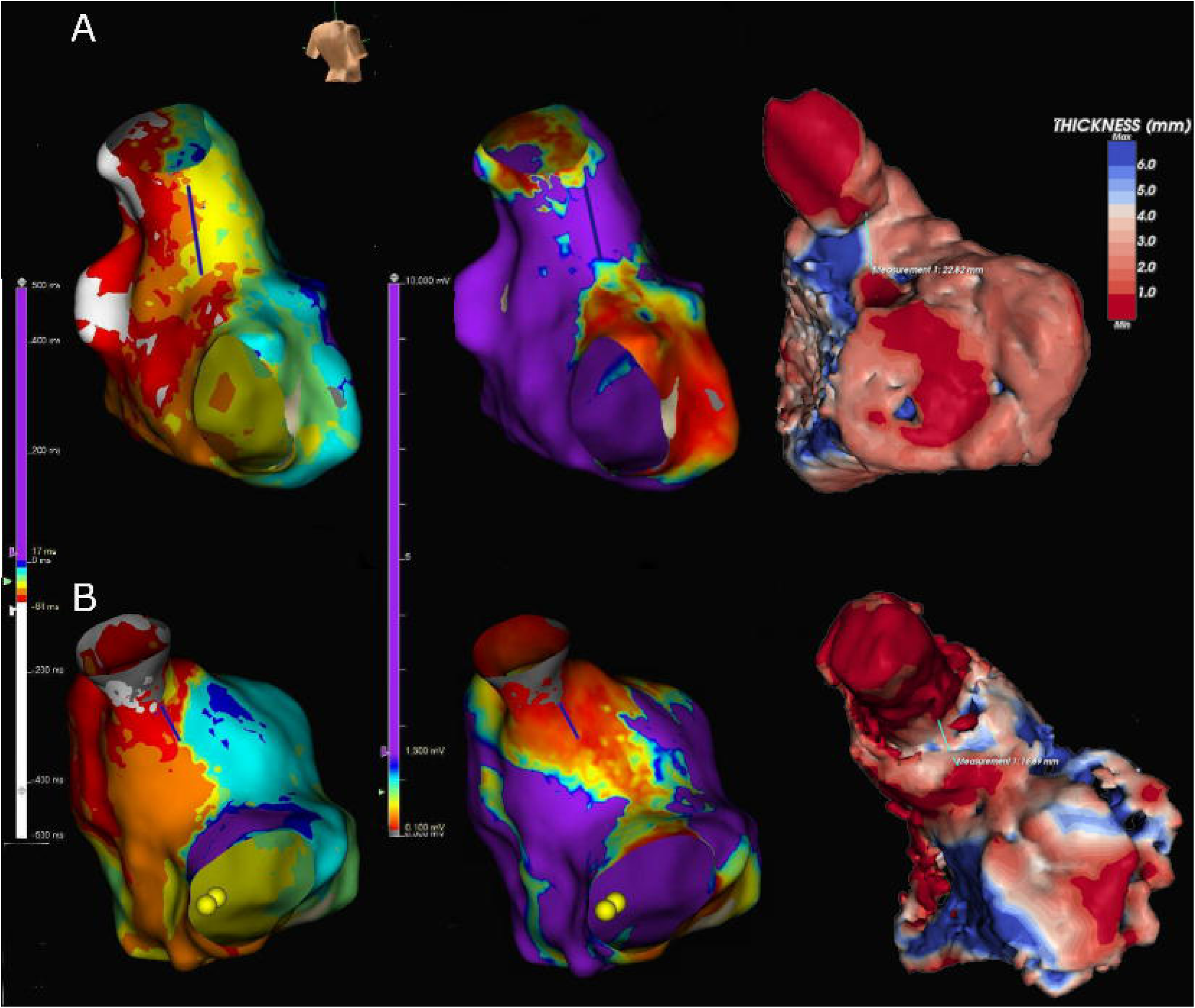
Representative examples of anatomic isthmus 3 in a patient without (A) and with (B) slow conducting anatomic isthmus 3 (SCAI 3). In both panels, the left image is an isochronal activation map (8 isochrones), the center image is an omnipolar voltage map with a dense scar defined as a voltage below 0.1 mV, and the right image is the parametric wall-thickness map of the three-dimensional reconstruction generated in ADAS 3D, in which color denotes wall thickness and tissue 1 mm thick or less is shown in deep red. The blue line marks anatomic isthmus 3 (AI 3). (A) Patient with normal conduction across AI 3. There is no isochronal crowding at AI 3, and the CT-derived AI 3 width is 22.8 mm. (B) Patient with a slowly conducting AI 3. Isochronal crowding is present at AI 3, corresponding to the low-voltage corridor on the omnipolar voltage map and to the narrower isthmus on CT, which measures 16.9 mm. AI 3 = anatomic isthmus 3; CT = computed tomography.

When focusing on the ten patients with SCAI 3 inducible VT, they also had significantly narrower isthmuses compared to patients without VT inducibility (18.6 ± 6.6 vs. 27.8 ± 9.5, p = 0.007). In univariate logistic regression, each 5 mm decrease in width was associated with 2.23-fold higher odds of inducible SCAI 3-dependent VT (95% CI 1.11–4.47, p = 0.022).

### AI 3 Width Measurement Was Reproducible

Inter-reader reproducibility was assessed in a subset of 10 patients. Five patients had SCAI 3, four of whom had SCAI 3 VT, and the remaining had normal conducting AI3. There were no differences between the subset and the rest of the population regarding sex, age at the time of study, and AI 3 length. Agreement between the two readers was good to excellent (ICC 0.91, 95% CI 0.71–0.98). Bland–Altmann analysis showed a mean bias of-1.19 mm with 95% limits of agreement of-8.9 to 6.6 mm, without evidence of proportional bias (Bland-Altmann regression slope= 0.197 (SE 0.134, 95% CI-0.112 to 0.506), p= 0.180, **Figure 3**)

**Figure 3.**
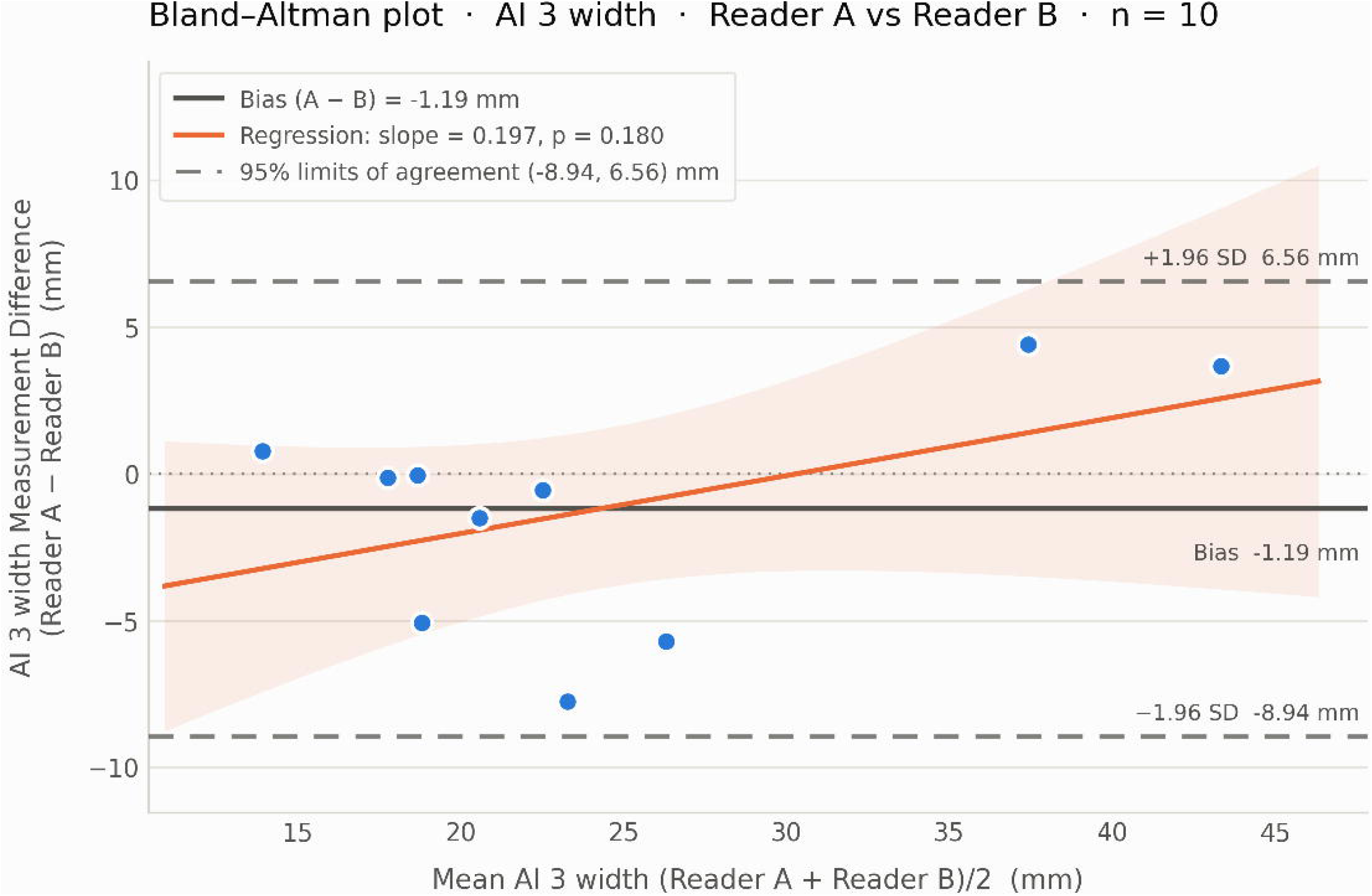
Bland Altmann plot for CT segmentation Assessment of inter-reader variability in the calculation of CT-derived Anatomic Isthmus 3 width. Agreement between the two readers was good to excellent (ICC 0.91, 95% CI 0.71–0.98; two-way random effects, absolute agreement, single measurement). Bland– Altmann analysis showed a mean bias of-1.14 mm with 95% limits of agreement of-8.9 to 6.6 mm, without evidence of proportional bias (Bland-Altmann regression slope= 0.197 (SE 0.134, 95% CI-0.112 to 0.506), p= 0.212). The orange line demonstrates the inter-reader regression, and the solid black line demonstrates the inter-reader bias. The light orange band is the 95% confidence interval of the proportional bias.

### Clinical Follow-up

All 12 patients with SCAI 3 underwent ablation. Two patients underwent additional ablation of a non-SCAI 3 isthmus-dependent VT (1 SCAI 2-dependent, 1 SCAI 1-dependent). The acute procedural endpoint of non-inducibility and bidirectional block across SCAI 3 was achieved in nine of the 12 ablated patients. For the three patients where the acute procedural endpoint was not met: (a) one had a deep intramural substrate in the LV summit that radiofrequency energy could not reach, (b) one had extension of the VT circuit to the right sinus of Valsalva in a significantly dilated aortic root, for which a retrograde aortic approach was deferred, and (c) one had an epicardial origin. Four patients underwent ICD implantation (two with and two without SCAI 3-dependent VT) as they were considered high risk of sudden cardiac death, not mitigated by the ablation procedure. The two patients who did not have SCAI 3-dependent VT had ICD implantation for secondary prevention due to hospital presentation with sustained monomorphic VT. Over a median follow-up of 6 months (IQR 125–365 days) after the index EPS, there were no deaths, no appropriate device therapies, and no repeat VT ablations.

### Age and Time Since Repair are Associated with AI 3 Width and VT Inducibility

Older age at EPS was associated with inducible SCAI 3-dependent VT (OR 2.13 per 10-year increase, 95% CI 1.07–4.27; p = 0.032). Similarly, a longer time since surgical repair was associated with higher odds for inducible SCAI 3-dependent VT (OR 2.69 per 10-year increase, 95% CI 1.15–6.22; p = 0.021). Examining the year of ToF surgery (with the first operation done in 1961), there was an inverse relationship between the probability of SCAI 3-dependent VT and time since surgery (OR 0.39 per 10-year interval from index surgery, 95% CI 0.17–0.89; p = 0.026). The AI 3 width was also inversely related to the time since surgical repair: each 10-year increase was associated with a 3.3 mm narrower isthmus (95% CI 0.5 – 6.3; R² = 0.15; p = 0.022; Figure 4).

## Discussion

In this study, we show that (a) the width of anatomic isthmus 3 measured on pre-procedural cardiac CT is narrower in patients with inducible SCAI 3-dependent VT than in those without; (b) narrower AI 3 width is associated with higher odds of SCAI 3 identification, and a threshold of 21.5 mm yields a negative predictive value of 80% and an accuracy of 77.8%; (c) the measurement is reproducible between independent readers blinded to each other and to the results of the electrophysiology study; and (d) in exploratory analyses, AI 3 width is inversely related to time since surgical repair, older age at EPS, and age at repair.

These findings are significant for several reasons: First, they suggest a non-invasive risk stratification tool with a high negative predictive value. Hence, this informs whom to bring to the electrophysiology laboratory. Second, nearly every candidate for

PV replacement has an existing CT scan so this is a widely available tool; therefore, the marker adds no additional test, radiation, or visit.. Third, they provide insight into substrate evolution in rTOF, indicating that the substrate is not static: AI 3 width was inversely associated with time since surgical repair and the patients had a higher risk for SCAI-3 dependent VT.

The biological plausibility of these findings rests on three linked observations: (a) the tissue bounding AI 3 is resolvable on CT as wall thinning, (b) the distance between those borders can be measured non-invasively, and (c) that distance is a determinant of the conduction properties that render the isthmus arrhythmogenic. Autopsy studies from patients with ToF have defined the tissue characteristics of the VT propagating isthmuses. The myocardium harboring the surgical scars demonstrates endocardial fibroelastosis and increased interstitial fibrosis of the surviving myocardial bundles.^6^ The tissue around the ventriculotomies, and the VSD patch is densely scarred and does not conduct. These non-conducting regions form the boundaries of AI 3, and the distance between them defines the corridor of surviving excitable myocardium through which activation must propagate. Patients with SCAI 3 related VT tend to have narrower and longer isthmuses of slow conducting myocardium on electroanatomical mapping.^18^ Isthmus width is itself a determinant of conduction. A wavefront emerging through a narrow corridor is diffracted into a curved wavefront and propagates slower than a planar wavefront.^19^ Contrast-enhanced CT resolves myocardial wall thickness at near-submillimeter resolution, and regional wall thinning co-localizes with low-voltage scar and the abnormal electrograms that sustain reentry.^20,21^ This correspondence between CT wall thinning and electroanatomic scar has been used to identify VT ablation targets in ischemic cardiomyopathy.^22^.

Invasive electroanatomic mapping defines the same isthmus directly: the anatomic isthmus is bounded by unexcitable tissue (patch material, valve annuli, and dense fibrosis), identified as bipolar voltage below 0.5 mV with loss of capture at pacing output above 10 mA, and its width is measured between these borders.^23^ On electroanatomic mapping, isthmuses supporting VT were longer, narrower, and more slowly conducting than those that did not (length 22 ± 7 vs 16 ± 7 mm; width 20 ± 8 vs 28 ± 11 mm; conduction velocity index 0.36 vs 0.78 m/s), and a slowly conducting isthmus predicted isthmus-related VT with 93% sensitivity and 100% specificity.^18^ This narrower mapped width parallels the AI 3 width we measured on CT (18.4 vs 29.2 mm), and over a mean of 50 months, VT occurred only in patients with a residual slowly conducting isthmus.^18^ Slow conduction across the isthmus, at a conduction velocity below 0.5 m/s, defines the arrhythmogenic slowly conducting isthmus.^24^ Our CT wall-thickness border is the anatomic counterpart of this electrically defined boundary, and AI 3 width on CT is the imaging analog of the mapped isthmus width. The magnitude of our measurement tracks the invasive data: AI 3 widths on CT in patients with and without inducible VT (18.6 vs 27.8 mm) approximate the electroanatomically mapped isthmus widths in the same comparison (20 vs 28 mm).^18^ The essential advantage is that CT yields this information without an invasive electrophysiology study.

In our exploratory analysis, we identified an inverse relationship between isthmus width and patient age and a positive relationship between patient age and the probability of inducible SCAI 3 VT. This is in accordance with other cohorts showing that older age is associated with age-related conduction slowing, and a rising probability of inducible VT.^25,26^ In a multicenter cohort of 180 patients studied before transcatheter PV replacement, isthmus conduction velocity fell by 0.08 m/s per decade, and VT inducibility increased with age.^25^ Our data are consistent with the hypothesis of an age-related progression of the substrate, in which chronic right ventricular remodeling after repair narrows the anatomic isthmus and slows its conduction over time, raising the probability of inducible SCAI 3–dependent VT. In the absence of serial electrophysiologic data, an age-dependent remodeling process is hypothesized rather than demonstrated, and longitudinal intra-patient mapping would be required to establish it.

Our study has limitations. First, despite representing one of the largest single-center ToF cohorts in the United States, our sample size was modest (n = 27, with 12 SCAI 3 cases), precluding multivariable logistic regression. Accordingly, although discrimination was internally validated with bootstrap optimism correction, the 21.5 mm cut-point and its negative predictive value were derived within the study sample and require prospective external validation before clinical use. Second, the study endpoint was identification of SCAI 3 and inducibility of SCAI 3-dependent VT at programmed stimulation rather than spontaneous clinical VT or sudden death; inducibility is an established but imperfect surrogate, and the short follow-up (median 6 months) precludes linking AI 3 width to clinical arrhythmic events. Third, AI 3 width addresses only the SCAI 3–dependent substrate; a wide, reassuring measurement does not exclude arrhythmic risk from other anatomic isthmuses, focal mechanisms, or atrial arrhythmias, so it cannot serve as a stand-alone risk-stratification tool. Fourth, AI 3 width was measured at a single center on one CT platform and one post-processing software; therefore, the 21.5 mm threshold may not transfer directly to other scanners or segmentation tools; metal artifacts from right ventricular device leads can also degrade visualization of outflow-tract reconstruction in this population. Standardized acquisition and cross-platform calibration are needed before wider application. Finally, the retrospective design allowed uncontrolled confounding; in particular, antiarrhythmic therapy or prior ablation before the index study may have reduced inducibility and misclassified the substrate. Prospective, multicenter validation with standardized CT acquisition and post-processing, incorporating conduction and functional measures and linked to clinical arrhythmic outcomes, is a necessary next step.

## Conclusion

In adults with repaired tetralogy of Fallot referred for pulmonary valve replacement, a narrower anatomic isthmus 3 on cardiac CT was associated with inducibility of SCAI 3-dependent ventricular tachycardia. Pending prospective multicenter validation, this measurement may offer a non-invasive means to identify which patients warrant screening electrophysiology study while the substrate remains accessible to ablation.

## Clinical Perspectives

1. In patients with Tetralogy of Fallot, AI 3 is a frequent source for VT. In patients that have undergone transcatheter pulmonic valve replacement, the site is inaccessible for ablation. Therefore, patients often undergo an EP study for VT inducibility before valve replacement.
2. Identifying a non-invasive marker for slow conduction AI 3 would allow many patients to forego the risks of a procedure. AI 3 width can be calculated from the pre-procedural cardiac CT after segmentation of the right ventricular wall.
3. In our study, we demonstrate that a narrow AI 3 is associated with slow conduction and VT inducibility. Thus, pending external validation, our marker could help identify what patients would benefit from an EP study before transcatheter valve replacement.

## Funding statement

This research received no specific grants from any funding agency in the public, commercial, or not-for-profit sectors.

## Disclosures

KNA has received honoraria from Johnson & Johnson MedTech for fellows’ program development, Future Cardia for non-clinical product testing, and consulting fees from NAMSA; these relationships are unrelated to the content of this manuscript. None of these relationships are with the manufacturers of the imaging, mapping, or ablation systems used in this study. JC is a consultant for Johnson & Johns MedTech, Abbott and Boston Scientific. The remaining authors have no conflicts of interest to disclose.

## Data Availability

Because this study describes a rare clinical entity with low prevalence, the deidentified patient data are not publicly available. A limited study dataset can be available by request to the corresponding author, Konstantinos Aronis at.

---

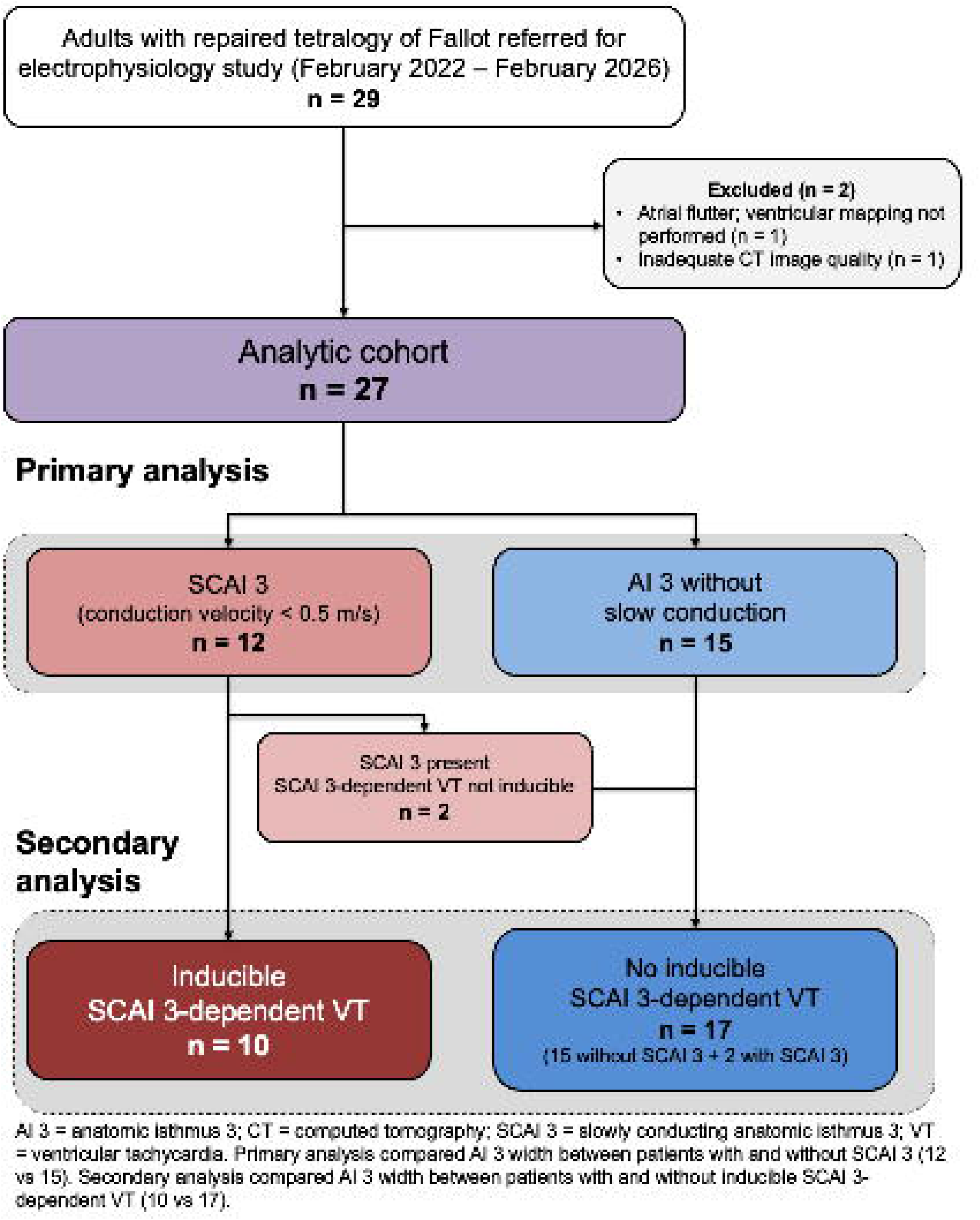

## Notes

### Author Declarations

Johns Hopkins Medicine Institutional Review Board gave ethical approval for this work.

