## Supplemental methods for "Narrow Is Slow: CT-Derived Anatomic Isthmus 3 Width and Arrhythmogenic Substrate in Repaired Tetralogy of Fallot"

***Supplemental Material, Methods and Results***

***for***

**Sizing the Substrate: CT-Derived Anatomic Isthmus 3 Width and Ventricular Tachycardia Inducibility in Repaired Tetralogy of Fallot**

***by***

Marinos, Kosmopoulos, MD, ^1,2^**^†^** Steven, Liskov, MD,^1,3^**^†^** Rawan Amir, MD, ^2,4^ Ronald, D, Berger, MD PhD^1^, Ari, Cedars, MD,^2,5^ Omar, Chehab, MD,^2,6^ Jonathan, Chrispin, MD^1^, Stacy, Fisher, MD, ^2,5^ Stephanie, Fuller, MD,^7^Hannah, Fraint, MD,^5^ Aimee, K, Armstrong, ^5^ MD,Ilias, Ziakas, MD,^1^Konstantinos, N, Aronis, MD, PhD^1,2^

#To whom correspondence should be addressed

**1. Supplemental Methods**

***1.1. Assessment of the Wall Thickness Threshold Defining the AI 3 Borders***

The parametric right ventricular (RV) wall thickness map generated in ADAS 3D (version 2; ADAS3D Medical, Barcelona, Spain) was exported for the first 14 consecutive patients, together with the corresponding map restricted to the segmented anatomic isthmus 3 (AI 3). Thickness was recorded at every vertex of the reconstructed surface, yielding a median of 14,155 RV vertices and 221 AI 3 vertices per patient. The exported thickness scalar was discretized by the software into 0.5-mm intervals with bin centers at 0.25 through 4.75 mm and saturation at the upper bin; values were assigned to these intervals for all patients so that distributions were directly comparable.

For each patient, we computed the median thickness and the proportion of vertices at or below candidate thresholds, separately for the entire RV and for AI 3. Patient-level values, rather than pooled vertices, were used for all statistical comparisons because vertices within a patient are spatially autocorrelated and are not independent observations. AI 3 and RV thickness were compared within patients using the Wilcoxon signed-rank test and between patients with and without inducible SCAI 3-dependent VT using the Wilcoxon rank-sum test. Analyses were performed in Python 3 (SciPy 1.1). This analysis was descriptive and was not corrected for multiple comparisons.

To examine whether the threshold could be derived from the data, we analyzed the underlying continuous thickness scalar, which resolves the structure below the 0.5-mm display interval. This continuous scalar was available in 16 of the 27 patients (due to software update mid-way our study). For each patient, we estimated the distribution of RV surface thickness by kernel density estimation and tested it for unimodality with the Hartigan dip test; the location of any minimum between the near-zero and myocardial portions of the distribution was recorded per patient and compared with the 1-mm rule.

***1.2. Exploratory Analysis: Unadjusted and Adjusted Associations Between Tachycardia Cycle Length, Anatomic Isthmus Width, and Age***

The analysis included patients with induced, mapped SCAI 3-dependent VT and complete data for AI 3 width, age at electrophysiology study, and VT cycle length (n = 10). VT cycle length was modeled by linear regression on age (cycle length ~ age), on AI 3 width (cycle length ~ width), and on both (cycle length ~ age + width). Spearman rank correlation was reported as a distribution-free check, collinearity was assessed by the variance inflation factor, and influence by Cook's distance. A prespecified sensitivity analysis excluded a single influential observation. Coefficient stability was tested with HC3 heteroscedasticity-consistent standard errors. The age–width association was estimated separately in the full cohort with both variables (n = 27). Adjusted associations were additionally expressed per interquartile range (IQR) of each predictor (AI 3 width, 5.65 mm; age, 10 years) to place them on a common scale, and the model-predicted cycle length was displayed as a fitted surface over the observed ranges of age and AI 3 width, with the convex hull of the analyzed observations demarcated to distinguish interpolation from extrapolation (Supplemental Figure S4).

***1.3 Study Covariates***

Baseline demographic, surgical, electrocardiographic, and echocardiographic covariates were extracted from the electronic medical records. Demographic and anthropometric covariates included age at EPS, sex, and body mass index. Surgical history covariates included age at ToF repair, number of prior thoracotomies, and interval since surgical repair. Arrhythmia history covariates included prior history of sustained VT and prior ICD implantation. Electrocardiographic covariates were derived from the 12-lead ECG recorded on the day of the EPS and comprised QRS duration, right bundle branch block, and QRS fragmentation; QRS fragmentation was classified as severe when more than two notches unrelated to a bundle branch block were present in the QRS in more than five leads[19]. Echocardiographic covariates were assessed on transthoracic echocardiography performed within one year of the procedure and comprised RV systolic function, RV basal diameter, and estimated RV systolic pressure. [

**2. Supplemental Results**

***2.1. Distribution of Right Ventricular Wall Thickness***

Wall thickness was measured at 196,429 RV vertices and 3,687 AI 3 vertices across the 14 patients (seven with and seven without inducible SCAI 3-dependent VT). AI 3 was thinner than the right ventricle as a whole in a paired within-patient comparison (median 2.75 mm versus 3.25 mm; median within-patient difference −0.50 mm, IQR −0.98 to −0.12; p = 0.014; Supplemental Figure S2A). The two distributions diverged principally between 1.5 and 3.0 mm, where AI 3 carried a greater proportion of its surface than the right ventricle, and above 4 mm, where it carried less (Supplemental Figure S2B). The median AI 3 thickness observed here is close to the median of 2.6 mm reported for anatomic isthmuses with documented conduction block on independent multidetector CT assessment and well below the 4.8 mm reported for isthmuses with slow conduction and 5.6 mm for those with normal conduction.[8]

A threshold of 1 mm excluded a mean of 4.1% of AI 3 vertices and 8.1% of RV vertices. The threshold coincides with the native boundary of the exported thickness map and can therefore be applied identically across studies without interpolation within a bin. Neither AI 3 thickness (median 2.66 mm versus 2.80 mm; p = 0.27), the proportion of AI 3 below 1 mm (p = 0.21), nor RV median thickness (p = 0.90) differed between patients with and without inducible SCAI 3-dependent VT. Thickness within the isthmus, therefore, did not distinguish the two groups in this subset, whereas isthmus width did in the full cohort.

***2.2. Structure of the wall-thickness distribution***

On the continuous maps (16 patients), the RV surface thickness distribution was bimodal rather than continuous, and unimodality was rejected in 14 of 16 patients (Hartigan dip test p < 0.05; Supplemental Figure S3A). A near-zero population, corresponding to the valve orifices, ventricular septal defect patch, and segmentation boundaries, accounted for a mean of 7.9% of the RV surface (range 0.9–22.5%) and was separated from the myocardial mode (approximately 2.0–3.3 mm) by a distinct minimum. This minimum was resolvable in 12 of 16 patients, at a median of 0.76 mm (interquartile range 0.47–0.82) and below 1 mm in 11 of these 12 patients (Supplemental Figure S3B, S3C). The 1-mm rule therefore lies immediately above the empirical minimum that separates the non-myocardial surface from the myocardial corridor.

***2.3. Justification of the 1-mm Border Threshold***

Taken together, these observations support the 1-mm threshold on three grounds. First, it is exactly representable. One millimeter coincides with a boundary between native intervals of the exported thickness map; therefore, the same rule can be applied to any study processed on this platform without interpolating within an interval, and it cannot drift between readers or between studies. Second, it is conservative. It excluded a mean of 4.1% of isthmus surface points and lies below the tenth percentile of AI 3 thickness, so it removes tissue at the margin rather than truncating the corridor itself. Because misclassification at this boundary widens rather than narrows the measured isthmus, it biases the association between width and inducibility toward the null. Third, it lies well below the thickness at which conduction block has been observed on independent multidetector CT assessment; therefore, tissue excluded by this rule is unlikely to have been conducting.

The selection of the threshold used no information on VT inducibility, and the threshold was not adjusted after the outcomes were known. We did not optimize the threshold against the inducibility outcome. The raw distribution contains a separable near-zero population, but it is non-myocardial, and its boundary with the myocardial corridor falls below 1 mm (median 0.76 mm); the 1-mm rule sits just above this boundary and at a native interval edge, so it was fixed on a priori grounds and applied unchanged to every measurement, rather than tuned within the sample.

**3. Supplemental Discussion**

***3.1. Associations Between Tachycardia Cycle Length, Anatomic Isthmus Width, and Age: Interpretation and Comparison with Prior Studies (Hypothesis Generating)***

The VT cycle length approximates the integral of the reciprocal of conduction velocity along the circuit and therefore depends on (a) effective path length, (b) conduction velocity, and (c) entrance and exit curvature. The CT-derived AI 3 width is a fixed anatomical distance between the pulmonary annulus, the VSD patch, and adjacent dense scar. Thus, it indexes only one dimension of the anatomic-corridor component; it is not equivalent to the complete reentry path or the functional isthmus delineated during tachycardia.

Framed this way, the finding is consistent with prior work once the compared quantities are distinguished. Experimental mapping across hearts has found that a larger isthmus length, width, narrowest width, and circuit path length are associated with a longer cycle length, directionally concordant with our results.[1] In repaired tetralogy of Fallot, slower isthmus conduction and impaired right ventricular function, not CT-derived AI 3 width, were correlated with longer cycle length, and that analysis was not restricted to AI 3.[2] In a high-resolution human mapping study, circuit dimensions did not distinguish fast from slow VT, while outer-loop conduction velocity was the consistent correlate of cycle length [PMID 33169628].[3] Our between-patient association may therefore identify a compact AI 3 substrate phenotype with a shorter transit distance where conduction does not slow proportionally, rather than evidence that narrowing accelerates conduction.

The direct analysis also addresses the role of age: within these patients, older age was associated with a longer cycle length, consistent with a prior report,[2] and the whole-cohort age–width association did not persist in the mapped subset; therefore, the finding does not imply that advancing age accelerates VT. Age confounding, if operative, would bias toward a longer cycle length and could only attenuate the observed association, not generate it.

These analyses are exploratory. Limitations include the small number of mapped VTs, sensitivity to a single influential observation, the absence of conduction-velocity and outer-loop measurements needed to separate anatomy from function, a cross-sectional design that cannot establish longitudinal electrophysiologic remodeling, and conditioning on referral, inducibility, mapping feasibility, and adequate CT. Confirmation requires larger cohorts that pair CT-derived isthmus geometry with invasively measured conduction velocity and right ventricular function.

**Supplemental Figures**

**Supplemental Figure S1.** Participant flow. Twenty-nine adults with repaired tetralogy of Fallot were referred for electrophysiology study between February 2022 and February 2026. Two patients were excluded: one was found to have atrial flutter and did not undergo ventricular mapping, and the other had inadequate right ventricular opacification on pre-procedural cardiac computed tomography angiography. The analytic cohort comprised 27 patients, each with a measurable anatomic isthmus 3 (AI 3) width on pre-procedural CTA and a measured AI 3 conduction velocity at electroanatomic mapping. In the primary analysis, patients were grouped by the presence (n = 12) or absence (n = 15) of slow conduction across AI 3 (SCAI 3, defined as a conduction velocity below 0.5 m/s). In the secondary analysis, patients were grouped by the inducibility of SCAI 3-dependent ventricular tachycardia at programmed ventricular stimulation.

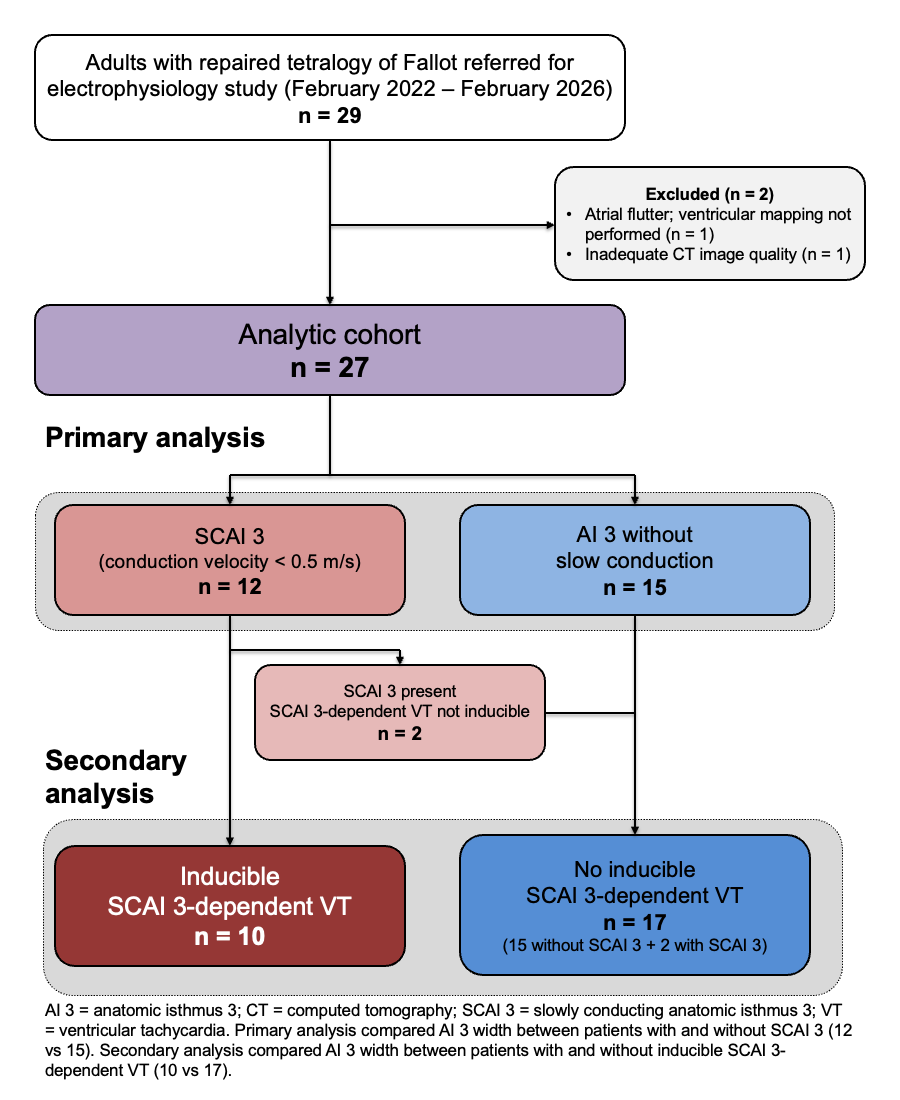

**Supplemental Figure S2.** Right ventricular and anatomic isthmus 3 wall thickness in the first 14 consecutive patients. (A) Median wall thickness within the entire right ventricle and within AI 3, paired by patient; horizontal bars denote group medians, and thin lines connect individual patients. (B) Distribution of wall thickness across the native 0.5-mm intervals of the exported thickness map, expressed as the mean proportion of surface points per patient; the error bars represent the standard errors across patients. The dashed line marks the 1-mm threshold used to define the isthmus borders, which coincided with the interval boundary. AI 3: anatomic isthmus 3; RV, right ventricle.

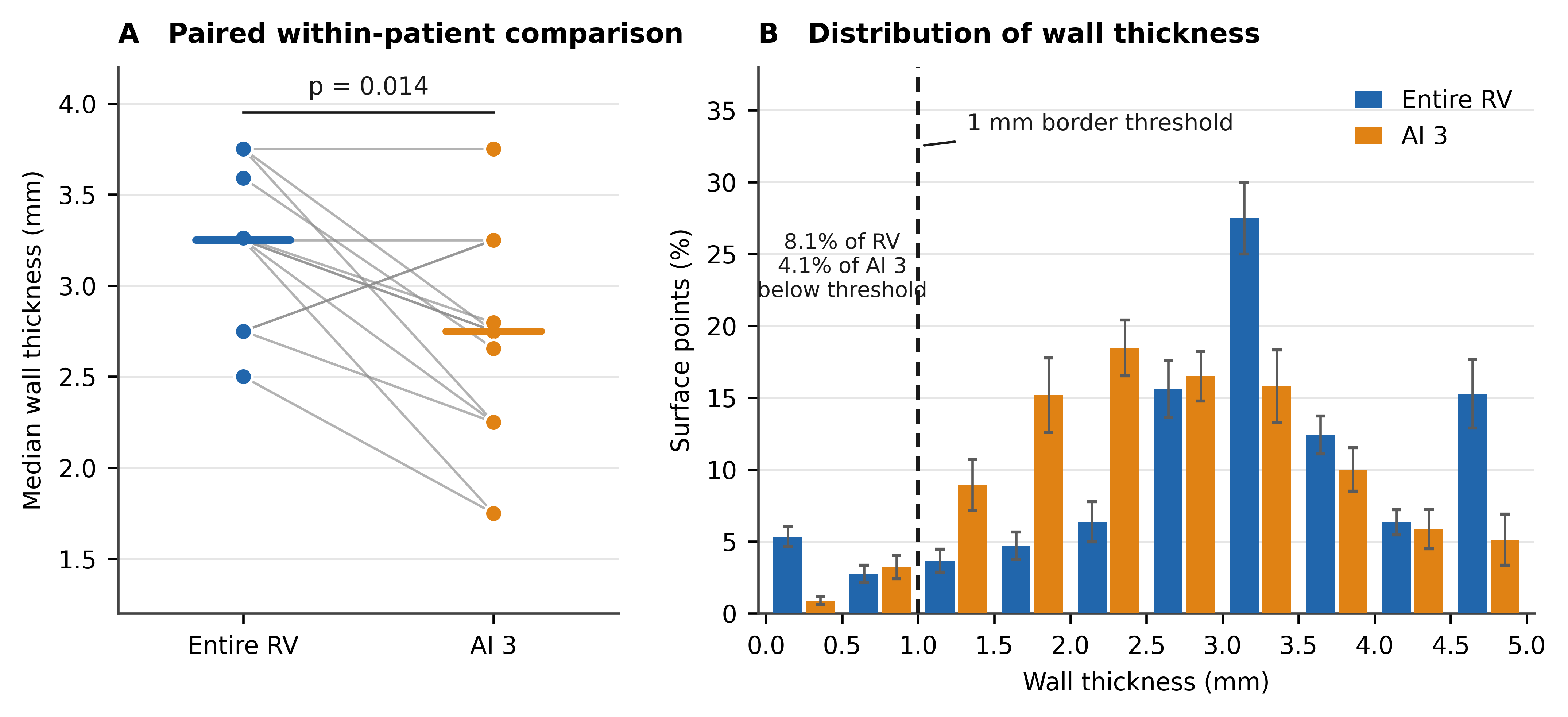

**Supplemental Figure S3.** Right ventricular wall-thickness distribution and the 1-mm isthmus-border threshold. Continuous (pre-discretization) RV wall-thickness maps were available in 16 of 27 patients. (A) Pooled distribution of surface thickness. The distribution is bimodal: a near-zero population (mean 7.9% of surface vertices), corresponding to valve orifices, the ventricular septal defect patch, and segmentation boundaries, is separated from the myocardial mode (~2.0–3.3 mm) by an antimode at 0.5–0.9 mm (shaded). The 1-mm border rule (red line) lies immediately above this antimode. (B) Per-patient kernel densities (0–3 mm); in most patients, a minimum precedes the myocardial rise, within the shaded band and below 1 mm. (C) Location of the per-patient thickness minimum (green) relative to the 1-mm rule (red) and the cohort median minimum (0.76 mm, dashed); × denotes patients in whom no distinct minimum was resolvable. AI 3: anatomic isthmus 3; RV: right ventricle.

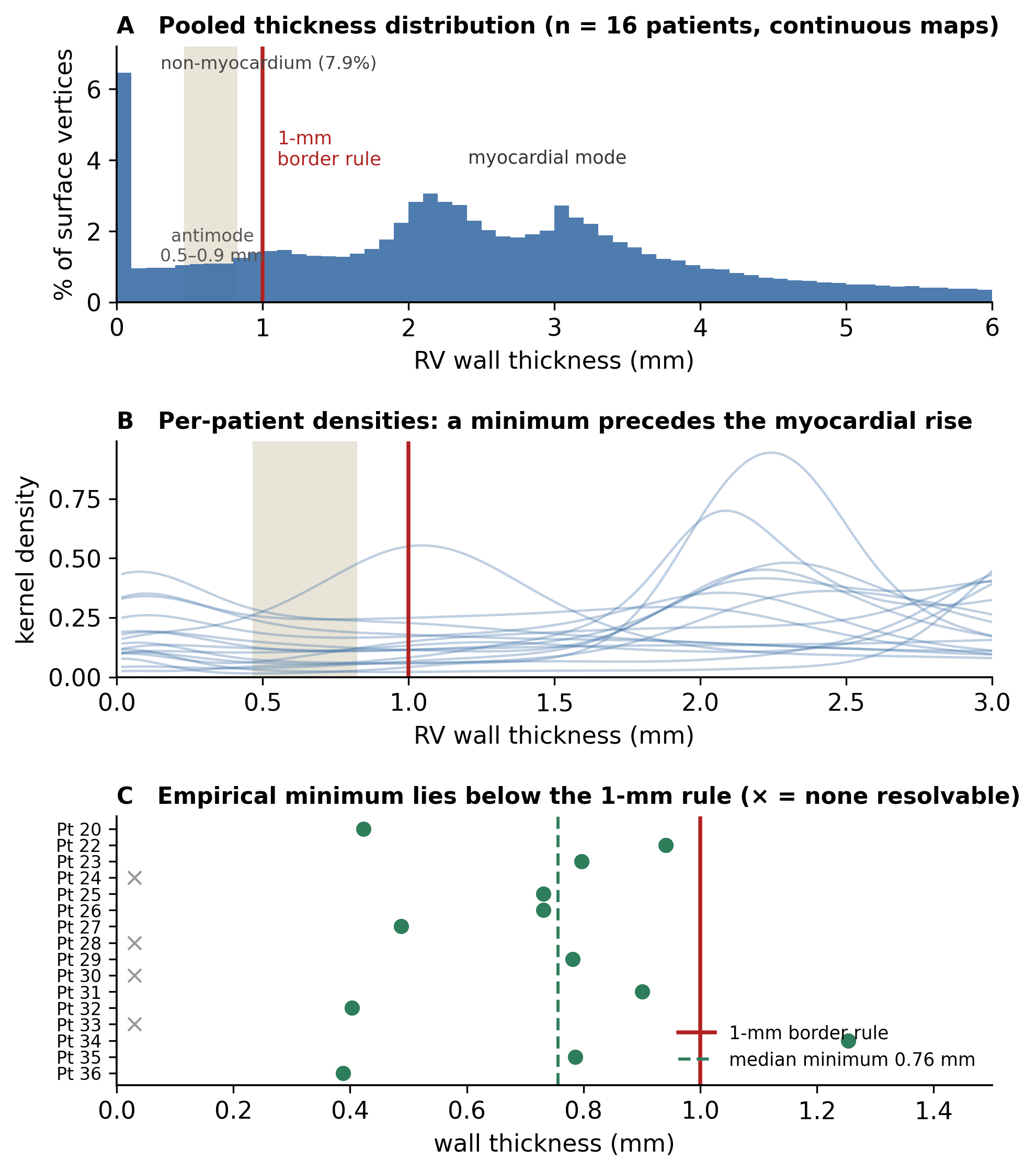

**Supplemental Figure S4.** Adjusted associations of anatomic isthmus 3 width and age with ventricular tachycardia cycle length in adults with repaired tetralogy of Fallot. **(A)** The colored surface and iso-TCL contour lines show predictions from the multivariable linear model fitted to nine mapped AI3-dependent VTs after exclusion of the influential observation: TCL (ms) = 33.58 + 0.884 × age (years) + 11.601 × AI3 width (mm). Hollow circles mark the age and AI3-width coordinates of the included VTs; the orange X identifies the influential observation, which was excluded from model estimation. Opaque color and solid contours represent interpolation within the convex hull of the included observations. Faded color and dashed contours represent extrapolation of the fitted linear model outside the convex hull but within the observed marginal ranges of age (28–62 years) and AI3 width (8.71–27.16 mm). No modeled surface is shown between the upper limit of the included width range and the excluded observation. The axes are scaled so that one observed IQR occupies the same physical distance in either direction (AI3-width IQR, 5.65 mm; age IQR, 10 years). On this scale, the near-vertical iso-TCL contours indicate that predicted TCL changes substantially more across AI3 width than across age. **(B)** IQR-scaled adjusted associations from the same multivariable model. Points represent model estimates, horizontal bars represent 95% CIs, and the dashed vertical line denotes no association. A 1-IQR greater AI3 width was associated with a 65.5-ms longer TCL (95% CI, 11.8–119.3 ms; P = .024), whereas a 1-IQR greater age was associated with an 8.8-ms longer TCL (95% CI, −38.5 to 56.2 ms; P = .664). These exploratory estimates describe adjusted associations and should not be interpreted causally. AI3 indicates anatomic isthmus 3; CI, confidence interval; EP, electrophysiology; IQR, interquartile range; TCL, tachycardia cycle length; and VT, ventricular tachycardia.

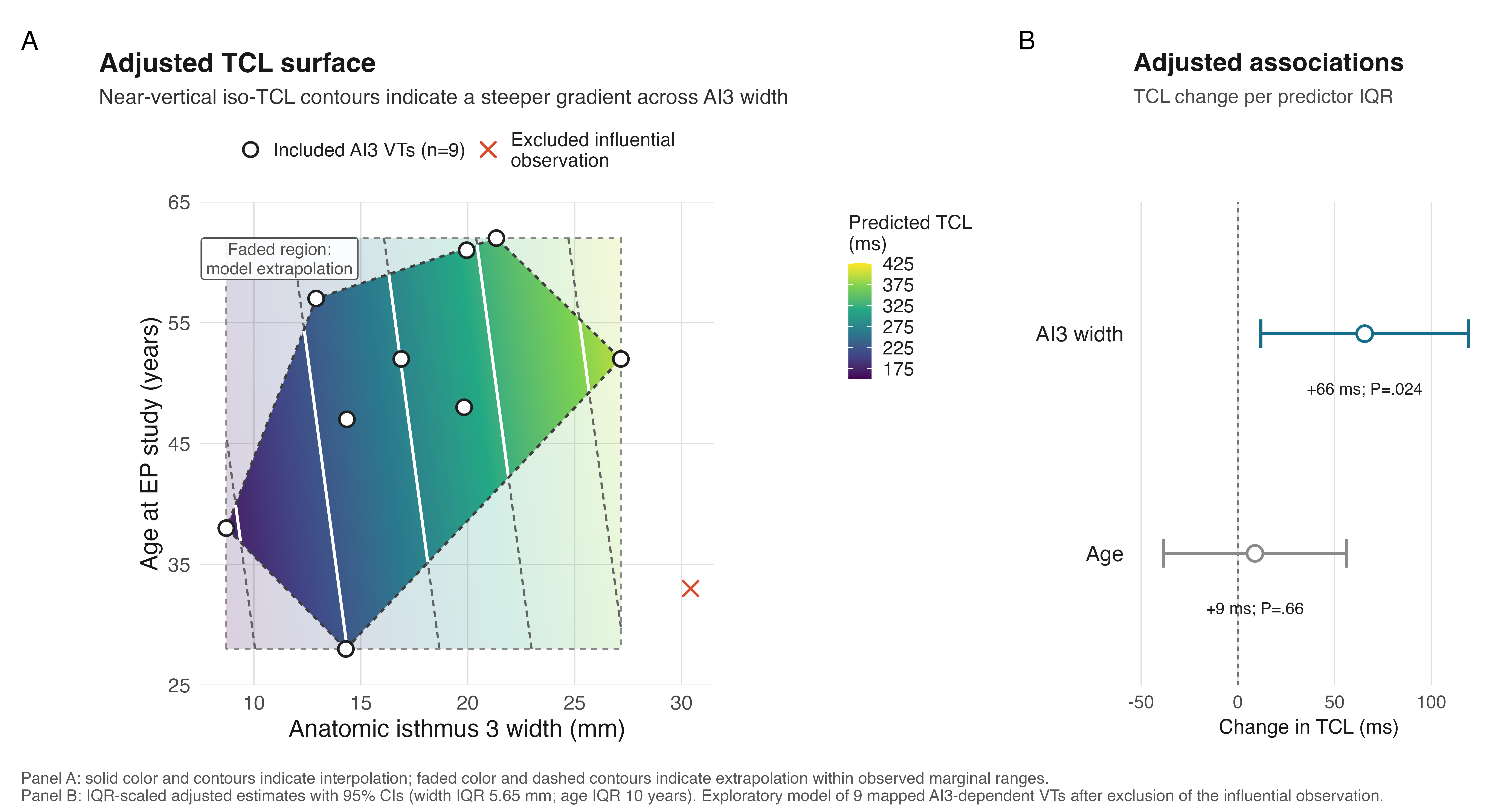
